# Trends in Awareness, Antihypertensive Medication Use and Blood Pressure Control by Race and Ethnicity Among Adults with Hypertension in the United States: A National Health and Nutrition Examination Analysis from 2011 to 2018

**DOI:** 10.1101/2021.08.18.21262251

**Authors:** Yuan Lu, Yuntian Liu, Lovedeep Singh Dhingra, Daisy Massey, César Caraballo, Shiwani Mahajan, Erica S. Spatz, Oyere Onuma, Jeph Herrin, Harlan M. Krumholz

**Affiliations:** Center for Outcomes Research and Evaluation, Yale New Haven Hospital, New Haven, Connecticut, USA; Section of Cardiovascular Medicine, Department of Internal Medicine, Yale School of Medicine, New Haven, Connecticut, USA; Department of Health Policy and Management, Yale School of Public Health, New Haven, Connecticut, USA

## Abstract

**Objective:** To examine United States (US) trends in racial/ethnic differences in hypertension awareness and antihypertensive medication use, and whether these differences help explain racial/ethnic differences in hypertension control among adults from 2011 to 2018.

**Design:** Population-based study.

**Setting:** National Health and Nutrition Examination Survey (NHANES), 2011-2018.

**Participants:** A nationally representative sample of US adults aged 18 years or older.

**Main outcome measures:** Hypertension awareness was defined as self-reported physician diagnosis of hypertension. Antihypertensive treatment was based on self-reported use of antihypertensive agents. Blood pressure (BP) control was measured systolic BP <140 mmHg and diastolic BP <90 mmHg.

**Results:** This study included 8,095 adults with hypertension from NHANES 2011-2018. During the study period, age-adjusted hypertension awareness declined from 84.0% (95% confidence interval 79.5 to 86.2) to 77.5% (74.0 to 80.5), hypertension treatment declined from 77.3% (73.4 to 81.1) to 71.4% (69.0 to 73.8) and control rates declined from 51.9% (47.1 to 56.7) to 43.1% (39.7 to 46.5). These declines were consistent for Black, Hispanic, and White individuals, but the three outcomes increased or did not change for Asian individuals. Compared with White individuals, Black individuals did not have a significantly different awareness (odds ratio:1.20 [0.96 to 1.45]) and overall treatment rate (1.04 [0.84 to 1.25]), but received more antihypertensive medications if treated (1.41 [1.27 to 1.56]), and had a lower BP control rate (0.72 [0.61 to 0.83]); Asian and Hispanic individuals had significantly lower awareness rates (0.69 [0.52 to 0.85] and 0.74 [0.59 to 0.89], respectively), overall treatment rates (0.72 [0.57 to 0.88] and 0.69 [0.55 to 0.82]), received fewer medications if treated (0.60 [0.50 to 0.72] and 0.86 [0.75 to 0.96]), and had lower BP control rates (0.66 [0.54 to 0.79] and 0.69 [0.57 to 0.81]). The racial/ethnic differences in awareness, treatment, and BP control persisted over the study period and were consistent across age, sex, and income strata. Lower awareness and treatment were associated with lower BP control in Asian and Hispanic individuals, but not in Black individuals.

**Conclusions:** Hypertension awareness, treatment, and control declined from 2011-2018, and this decline was consistent for Black, Hispanic, and White individuals. BP control was worse for Asian, Black, and Hispanic individuals than for White individuals over the entire study period; this was explained partly by differences in awareness and treatment for Asian and Hispanic individuals, but not for Black individuals.

**What this Paper Adds:** *Section 1: What is already known on this subject?:* Hypertension control rate has declined in the United States (US) since 2013, and this decline is more pronounced in Black and Hispanic communities.

*Section 2: What this study adds:* Our study shows that hypertension awareness, treatment, and control in the US have worsened from 2011 to 2018, and this trend was consistent for Black, Hispanic, and White individuals. Compared with White individuals, Black individuals had similar awareness and overall treatment rates, received more antihypertensive medications if treated, but had a lower control rate; Asian and Hispanic individuals had significantly lower awareness rates, overall treatment rates, received fewer medications if treated, and had lower control rates. Lower awareness and treatment were associated with lower BP control in Asian and Hispanic individuals, but not in Black individuals. These findings highlight the need for interventions to improve awareness and treatment among Asian and Hispanic individuals, and more investigation into the downstream factors that may contribute to the poor hypertension control among Black individuals.

## BACKGROUND

Progress in hypertension control in the United States (US) has been slow over the past decade. Despite ongoing efforts to improve hypertension control, the national blood pressure (BP) control rate (previously defined as BP <140/90 mmHg) among those with hypertension dropped from 54% in 2013-2014 to 44% in 2017-2018.^1^ The decline in BP control is more pronounced in Black and Hispanic communities.^1 2^ Identifying factors impeding BP control is necessary to inform future strategies to address race/ethnicity equity in cardiovascular diseases.

Lack of awareness and underuse of guideline to recommended medications are among the critical factors contributing to poor BP control.^3^ Prior studies show that Black individuals were more aware of their hypertension and were more likely to be on treatment compared with their White counterparts,^2 4 5^ while Hispanic individuals were more likely to be untreated or undertreated for hypertension.^5-7^ These studies, however, did not assess recent trends, nor did they investigate racial/ethnic differences in the intensity of treatment (number and type of medication), which may play an important role in the control of hypertension.^8 9^ Additionally, there are no recent studies of hypertension awareness, treatment, or control in the Asian American population, a group that is increasing in size and has high hypertension prevalence.^10 11^ Another knowledge gap is how much progress the US has made in eliminating disparities in awareness and treatment of hypertension and whether these differences in awareness and treatment sufficiently explain the difference in hypertension control.

Accordingly, we leveraged data from the National Health and Nutrition Examination Survey (NHANES),^12^ which provides estimates of a nationally representative sample of the US population, to evaluate racial/ethnic disparities in hypertension awareness, treatment, and control from 2011 to 2018. Due to the influence of income on health and health care access in the US,^13^ we also evaluated how racial/ethnic differences varied by income. Finally, we assessed whether racial/ethnic differences in awareness and treatment were associated with differences in hypertension control. By better understanding the racial/ethnic differences in hypertension management in the US, we sought to identify targets for public health interventions.

## METHODS

### Study design and population

We included data from 23,825 adults, aged ≥18 years, included in the NHANES for the years 2011–2018. The NHANES is a series of cross-sectional, weighted, multistage sampled surveys that provide nationally-representative estimates on the non-institutionalized US population.^12^ Since 1999, NHANES has been conducted in 2-year cycles. Survey participants received in-home interviews, followed by standardized physical examinations conducted in mobile examination centers, and laboratory tests using blood and urine specimens provided by participants during the physical examination. For the current analysis, we used data from 4 cycles conducted from 2011-2012 through 2017-2018 to focus on recent trends. During our study period, the mean participant response rate was 65.3% for interviews and 62.3% for physical examinations.^14^ We categorized individuals into four mutually exclusive subgroups based on their self-reported race/ethnicity information: non-Hispanic Asian, non-Hispanic Black, Hispanic, and non-Hispanic White subgroups. For simplicity, we hereafter refer to the study groups as Asian, Black, Hispanic, and White. We excluded individuals who identified as Alaskan Native or American Indian or ‘Other’ Race (n=876) due to small numbers. We also excluded pregnant women (n=179, Appendix Figure 1).

### Measurements of hypertension awareness, antihypertensive medication use, and BP

During the home interviews, participants were asked if they had ever been told by a doctor or other health professional that they had hypertension or high BP. Those who answered “yes” were then asked if they were currently taking prescribed medicine for hypertension. Those who answered “yes” were then further asked to show the interviewer the medication containers of all the products used in the past 30 days. For each medication reported, the interviewer entered the product’s complete name from the container into a computer. If no container was available, the interviewer asked the participant to verbally report the name of the medication.

During the standardized physical examinations, participants’ BP levels were measured by trained clinicians using a mercury sphygmomanometer and an appropriately sized BP cuff. Three consecutive BP measurements were obtained after participants rested quietly in a seated position for 5 minutes. If a BP measurement was interrupted or incomplete, a fourth attempt was made. Mean systolic and diastolic BP were calculated for each individual per NHANES reporting guidelines.^15^

### Definitions of hypertension, awareness, treatment, and control

Consistent with previous studies by Muntner et al and with the JNC 8 hypertension guidelines available during the time of this study,^1 16^ hypertension was defined as a mean systolic blood pressure (SBP) level of 140 mmHg or higher, a mean diastolic blood pressure (DBP) level of 90 mmHg or higher, or an affirmative response to “are you now taking prescribed medicine to lower your blood pressure?” Among those with hypertension, awareness was defined by an affirmative response to the question: “Have you ever been told by a doctor or other health care professional that you had hypertension, also called high BP?” Among those with hypertension, individuals who had the name of antihypertensive medication documented (either individuals showed the interviewer the medication containers or verbally reported the name of the medication, as described above) were categorized as taking antihypertensive medications. BP control was defined as a mean SBP level lower than 140 mmHg and a mean DBP level lower than 90 mmHg.

In the sensitivity analyses, we used thresholds from the 2017 American College of Cardiology/American Heart Association (ACC/AHA) BP guideline to define hypertension and BP control. Hypertension was defined as a mean SBP level of 130 mmHg or higher, a mean DBP level of 80 mmHg or higher, or self-reported antihypertensive medication use. BP control was defined as a mean SBP level lower than 130 mmHg and a mean DBP level lower than 80 mmHg.

### Class and number of antihypertensive medications

Antihypertensive medications were categorized into the following classes: (1) angiotensin-converting enzyme inhibitors (ACEIs), (2) angiotensin II receptor blockers (ARBs), (3) calcium channel blockers (CCBs), (4) β-blockers, (5) thiazide and thiazide-like diuretics, and (6) others (e.g., other diuretics, direct vasodilators, renin inhibitors, α1-blockers, and other centrally acting drugs, see list of medications in Appendix Table 1). Monotherapy was defined when a person reported taking only one class of antihypertensive agents. Combination therapy was defined when a person reported taking more than one class of antihypertensive agents, including single-pill combination drugs.

### Other sociodemographic, behavioral, and clinical variables

We included other variables in the analysis, including age (in years), sex (male, female), education level (less than high school, high school diploma, some college, Bachelor’s degree or higher), family income (based on the percent of family income relative to the federal poverty limit from the Census Bureau: high/middle income [≥200%] and low-income [<200%]), insurance status (insured, uninsured), marital status (married, unmarried), employment status (working, not in the labor force, unemployed), smoking status (current, former, never smoker), alcohol intake (never, former, light drinker, moderate drinker, heavy drinker), physical activity (recommended, inactive, insufficient), obesity defined as body mass index (BMI) ≥ 30kg/m^2^, diabetes defined as previous physician diagnosis or HbA1c ≥ 6.5% or currently on antidiabetic medication, self-reported history of hyperlipidemia, myocardial infarction (MI) or stroke, cancer, and kidney disease. Detailed definitions of the covariates are reported in Appendix Table 2. Information on all sociodemographic variables was available for all years and responses coded as “unknown” or “not ascertained” were analyzed under a separate category of “unknown.”

### Statistical analysis

All analyses used methods appropriate for structured survey data, incorporating strata and weights to produce nationally representative estimates. All person weights were pooled and divided by the number of years studied, following the NHANES guidance.^12^

We first described sociodemographic and clinical characteristics among hypertensive adults by race/ethnicity. Among all hypertensive individuals, we estimated the age-adjusted annual rates of awareness, overall treatment, and BP control by racial/ethnic subgroups using multivariable linear regression models. A separate model was estimated for each racial/ethnic subgroup and each outcome, including standardized age and an indicator for each survey year as independent variables. The coefficients for each year then represented the age-adjusted annual rates for the designated outcome.^17^ Using a similar approach, we estimated the age-adjusted annual rate for number and class of antihypertensive mediation use by racial/ethnic subgroup, among treated hypertensive individuals. The trend for each outcome was estimated by a weighted linear regression, using the reciprocal of the annual rate’s standard error as weights.

We estimated the odds ratios (ORs) for each outcome comparing each racial/ethnic group relative to their White counterparts using multivariable logistic regression models. We conducted two models: a minimally adjusted model adjusting for age (categorized as 18-39, 40-59, 60-79, 80+ years of age) and sex, and a fully adjusted model additionally adjusting for individuals’ sociodemographic, behavior, and clinical characteristics. These characteristics included family income, insurance status, smoking status, diabetes, kidney disease, and cardiovascular disease (MI or stroke), all of which have been shown to be relevant in previous studies to hypertension treatment and control. We reported the OR of both minimally adjusted and fully adjusted models.

Finally, we assessed how racial/ethnic differences in BP control changed after accounting for differences in hypertension awareness and treatment using a propensity score weighting method.^18 19^ Specifically, we applied gradient boosting models to estimate the propensity scores for predicting race/ethnicity. The covariates used in the propensity score estimation included age, sex, education level, marital status, family income, insurance status, employment status, smoking status, physical activity, BMI, and comorbidities (diabetes, hyperlipidemia, MI, stroke, kidney disease, cancer). Kolmogorov-Smirnov statistic and the corresponding P values were used to assess covariate balance after propensity score weighting. We then developed two weighted regression models to estimate the racial/ethnic differences in BP control rate – one included race/ethnicity as the independent variable and one additionally included hypertension awareness and antihypertension treatment. Survey weights were incorporated in both the process of the propensity scores estimation, and the racial/ethnic difference estimation.

We considered 2-sided P values <0.05 to be statistically significant. All analyses were performed using R 4.0. This study received an exemption for review from the Institutional Review Board at Yale University because NHANES data are publicly available and de-identified. The study was reported following the STROBE (Strengthening the Reporting of Observational Studies in Epidemiology) reporting guidelines.^20^

## RESULTS

A total of 22,770 adults were included in NHANES 2011-2018, of whom 8,095 had hypertension and were in the final analysis (Appendix Figure 1). Among individuals with hypertension, the mean age was 60.3 [SD, 13.8] years, 51.5% (95% confidence interval: 50.0 to 53.0) were women, 4.6% (3.7 to 5.5) were Asian, 15.0% (12.4 to 17.7) were Black, 10.9% (8.8 to 13.0) were Hispanic, and 69.5% (65.7 to 73.2) were White. Asian and White individuals were older, had a higher income level, were more likely to be physically active, and were more likely to have health insurance compared with Black and Hispanic individuals (Table 1). Among all four racial/ethnic groups, Asian individuals had the highest prevalence of obesity and diabetes and the lowest percentages of current smokers and moderate/heavy drinkers.

**Table 1.** General characteristics of hypertensive adults by race/ethnicity in 2011 to 2018

| % (95% CI) | Asian | Black | Hispanic | White |
| --- | --- | --- | --- | --- |
| **Total** | 803 (4.6%, [3.7 to 5.5]) | 2452 (15%, [12.4 to 17.7]) | 1700 (10.9%, [8.8 to 13]) | 3140 (69.5%, [65.7 to 73.2]) |
| **Age (years)** |  |  |  |  |
| 18-39 | 58 (9.9%, [7.2 to 12.6]) | 203 (12.8%, [10.8 to 14.8]) | 109 (13.3%, [10.5 to 16.1]) | 205 (7%, [5.7 to 8.3]) |
| 40-59 | 287 (36.2%, [32.8 to 39.6]) | 879 (45.5%, [42.7 to 48.3]) | 517 (44.2%, [40.7 to 47.8]) | 805 (33.9%, [31.9 to 35.8]) |
| 60-79 | 399 (46.3%, [42 to 50.6]) | 1199 (36%, [33.7 to 38.3]) | 960 (37.4%, [34.1 to 40.8]) | 1385 (46%, [43.8 to 48.2]) |
| 80+ | 59 (7.6%, [5.3 to 10]) | 171 (5.7%, [4.6 to 6.8]) | 114 (5%, [3.8 to 6.2]) | 745 (13.1%, [11.8 to 14.5]) |
| **Sex** |  |  |  |  |
| Women | 396 (50.4%, [47.4 to 53.3]) | 1275 (56.7%, [54.7 to 58.7]) | 922 (51.1%, [48.1 to 54.2]) | 1555 (50.6%, [48.6 to 52.6]) |
| Men | 407 (49.6%, [46.7 to 52.6]) | 1177 (43.3%, [41.3 to 45.3]) | 778 (48.9%, [45.8 to 51.9]) | 1585 (49.4%, [47.4 to 51.4]) |
| **Education level** |  |  |  |  |
| More than high school | 479 (59.9%, [54.8 to 65.1]) | 1173 (49.6%, [46.4 to 52.8]) | 520 (34.7%, [30.6 to 38.7]) | 1760 (62.6%, [59.5 to 65.6]) |
| High school | 132 (17.1%, [13.8 to 20.3]) | 670 (27.6%, [25.9 to 29.3]) | 312 (21%, [18.4 to 23.5]) | 850 (26.3%, [23.8 to 28.7]) |
| Less than high school | 192 (23%, [18.5 to 27.5]) | 603 (22.8%, [20.1 to 25.6]) | 863 (44.4%, [40 to 48.7]) | 527 (11.2%, [9.1 to 13.2]) |
| **Family income** |  |  |  |  |
| High/middle income | 442 (65.4%, [58.8 to 72.1]) | 1009 (48%, [44.4 to 51.7]) | 539 (40.9%, [36.9 to 45]) | 1583 (70.7%, [67.3 to 74.1]) |
| Low income | 252 (34.6%, [27.9 to 41.2]) | 1131 (52%, [48.3 to 55.6]) | 908 (59.1%, [55 to 63.1]) | 1326 (29.3%, [25.9 to 32.7]) |
| **Insurance status** |  |  |  |  |
| Uninsured | 86 (9.5%, [7.3 to 11.8]) | 295 (13.7%, [11.6 to 15.8]) | 303 (20.4%, [17.6 to 23.2]) | 231 (6.5%, [5 to 7.9]) |
| Insured | 717 (90.5%, [88.2 to 92.7]) | 2151 (86.3%, [84.2 to 88.4]) | 1395 (79.6%, [76.8 to 82.4]) | 2904 (93.5%, [92.1 to 95]) |
| **Marital status** |  |  |  |  |
| Married/Living with partner | 600 (74.2%, [70.3 to 78.1]) | 1075 (44.1%, [41.6 to 46.6]) | 1029 (62.3%, [59 to 65.6]) | 1823 (65.1%, [62.9 to 67.3]) |
| Unmarried | 202 (25.8%, [21.9 to 29.7]) | 1369 (55.9%, [53.4 to 58.4]) | 667 (37.7%, [34.4 to 41]) | 1314 (34.9%, [32.7 to 37.1]) |
| **Employment Status** |  |  |  |  |
| Not in labor force | 382 (47.3%, [42.4 to 52.2]) | 1260 (47.1%, [44.6 to 49.6]) | 921 (46.4%, [42.4 to 50.3]) | 2028 (53.3%, [51 to 55.6]) |
| Unemployed | 16 (2.1%, [0.8 to 3.4]) | 88 (4.2%, [3.4 to 5]) | 51 (3.9%, [2.8 to 4.9]) | 61 (2.1%, [1.2 to 3]) |
| With a job/Working | 387 (50.6%, [45.7 to 55.4]) | 1032 (48.7%, [46.2 to 51.2]) | 673 (49.8%, [46 to 53.5]) | 998 (44.6%, [42.3 to 46.9]) |
| **Smoking Status** |  |  |  |  |
| Current smoker | 66 (8.9%, [6.4 to 11.4]) | 619 (25.6%, [23.1 to 28.1]) | 225 (15.2%, [13 to 17.4]) | 582 (17.4%, [15.6 to 19.2]) |
| Former smoker | 157 (18.7%, [15.8 to 21.5]) | 584 (20.4%, [18.2 to 22.6]) | 486 (27.6%, [24.8 to 30.4]) | 1152 (34.8%, [32.4 to 37.2]) |
| Never smoker | 580 (72.4%, [68.8 to 76]) | 1246 (54%, [51.5 to 56.5]) | 987 (57.2%, [54.1 to 60.3]) | 1402 (47.8%, [45.4 to 50.2]) |
| **Physical activity** |  |  |  |  |
| Recommended | 249 (31.2%, [26.9 to 35.4]) | 617 (25.9%, [23.8 to 28.1]) | 353 (22.1%, [19.5 to 24.8]) | 767 (29.5%, [27.4 to 31.7]) |
| Inactive | 416 (51.4%, [47.4 to 55.3]) | 1472 (58.9%, [56.2 to 61.6]) | 1111 (63%, [59.6 to 66.3]) | 1938 (55%, [52.4 to 57.6]) |
| Insufficient | 137 (17.5%, [14.3 to 20.6]) | 362 (15.2%, [13.8 to 16.6]) | 234 (14.9%, [12.5 to 17.3]) | 430 (15.5%, [14.1 to 16.9]) |
| **Alcohol intake** |  |  |  |  |
| Never | 226 (34.3%, [29.7 to 38.9]) | 300 (14.7%, [13.1 to 16.2]) | 324 (18.9%, [16.4 to 21.4]) | 297 (9.6%, [8 to 11.2]) |
| Former | 118 (18%, [14.3 to 21.7]) | 606 (25.7%, [23.4 to 28]) | 360 (21.5%, [18.8 to 24.3]) | 745 (21.9%, [20.2 to 23.6]) |
| Light drinker | 254 (39.1%, [34.2 to 43.9]) | 864 (44.2%, [41.6 to 46.8]) | 592 (41.8%, [39 to 44.7]) | 1160 (46.5%, [43.8 to 49.3]) |
| Moderate drinker | 38 (6.2%, [3.6 to 8.7]) | 212 (9.8%, [8.6 to 10.9]) | 152 (12.8%, [10.9 to 14.7]) | 326 (13.3%, [11.8 to 14.9]) |
| Heavy drinker | 14 (2.4%, [1 to 3.7]) | 114 (5.6%, [4.4 to 6.8]) | 55 (4.9%, [3.3 to 6.5]) | 192 (8.6%, [7.2 to 10]) |
| **BMI, kg/m2** |  |  |  |  |
| <25 | 317 (40.5%, [36.8 to 44.2]) | 427 (16.6%, [14.9 to 18.3]) | 180 (9.6%, [7.9 to 11.2]) | 590 (17.5%, [15.8 to 19.1]) |
| 25 to <30 | 134 (19.1%, [16.6 to 21.7]) | 1293 (58.1%, [55.9 to 60.3]) | 888 (57.2%, [54 to 60.4]) | 1423 (50.4%, [47.9 to 52.9]) |
| >=30 | 314 (40.4%, [36.3 to 44.4]) | 626 (25.3%, [23.6 to 27]) | 548 (33.2%, [30.5 to 36]) | 953 (32.1%, [30.1 to 34.2]) |
| **Comorbidities** |  |  |  |  |
| Diabetes | 283 (34.2%, [30.3 to 38.2]) | 836 (31.2%, [29 to 33.3]) | 652 (34%, [30.9 to 37.1]) | 822 (24.4%, [22.8 to 26]) |
| Hyperlipidemia | 408 (49.2%, [45.4 to 53.0]) | 1205 (48.1%, [45.7 to 50.5]) | 924 (50.1%, [47.2 to 53.1]) | 1774 (56.3%, [53.8 to 58.9]) |
| Kidney disease | 36 (4.4%, [2.8 to 6]) | 192 (7.4%, [6.3 to 8.5]) | 135 (6.8%, [5.6 to 8]) | 233 (5.6%, [4.6 to 6.6]) |
| Heart attack | 31 (3.4%, [2.2 to 4.7]) | 155 (5.4%, [4.5 to 6.3]) | 132 (6.4%, [5 to 7.8]) | 362 (8.2%, [7.1 to 9.2]) |
| Heart failure | 19 (2%, [1.1 to 2.9]) | 181 (6.9%, [5.5 to 8.3]) | 108 (5.2%, [3.8 to 6.6]) | 293 (6.1%, [5.1 to 7.2]) |
| Stroke | 34 (4.2%, [2.8 to 5.6]) | 218 (7.6%, [6.5 to 8.8]) | 97 (4.9%, [3.8 to 6.1]) | 296 (6.7%, [5.9 to 7.5]) |
| Cancer | 49 (5.9%, [4.4 to 7.4]) | 249 (8.5%, [7.4 to 9.6]) | 155 (7.2%, [5.9 to 8.6]) | 791 (22.8%, [21 to 24.6]) |
| **SBP, mean (95% CI), mmHg** | 139.9 (139.5 to 141.2) | 140.3 (139.3 to 141.3) | 139.3 (137.9 to 140.6) | 135.7 (134.6 to 136.7) |
| **DBP**, mean (95% CI), mmHg | 77.3 (76.2 to 78.4) | 77.1 (76.0 to 78.1) | 76.5 (75.2 to 77.9) | 73.5 (72.6 to 74.3) |
| Abbreviations: CI, confidence interval; BMI, body mass index; SBP, systolic blood pressure; DBP, diastolic blood pressure. |  |  |  |  |

### Trends in hypertension awareness by race and ethnicity

From 2011-2012 to 2017-2018, the age-adjusted hypertension awareness rate declined from 84.0% (79.5 to 86.2) to 77.5% (74.0 to 80.5) in the overall population. This decline was consistent for Black (86.4% [83.9 to 88.9] vs. 82.6% [78.5 to 86.7]), Hispanic (82.2% [78.1 to 86.2] vs. 73.6% [68.5 to 78.7]), and White (82.9% [78.2 to 87.6] vs. 77.4% [72.4 to 82.3]) individuals. However, hypertension awareness rate did not change for Asian individuals (72.6% (67.1 to 78.1) vs. 78.0% [70.4 to 85.7]) although their initial awareness rates were significantly lower than other groups (Figure 1).

**Figure 1.**
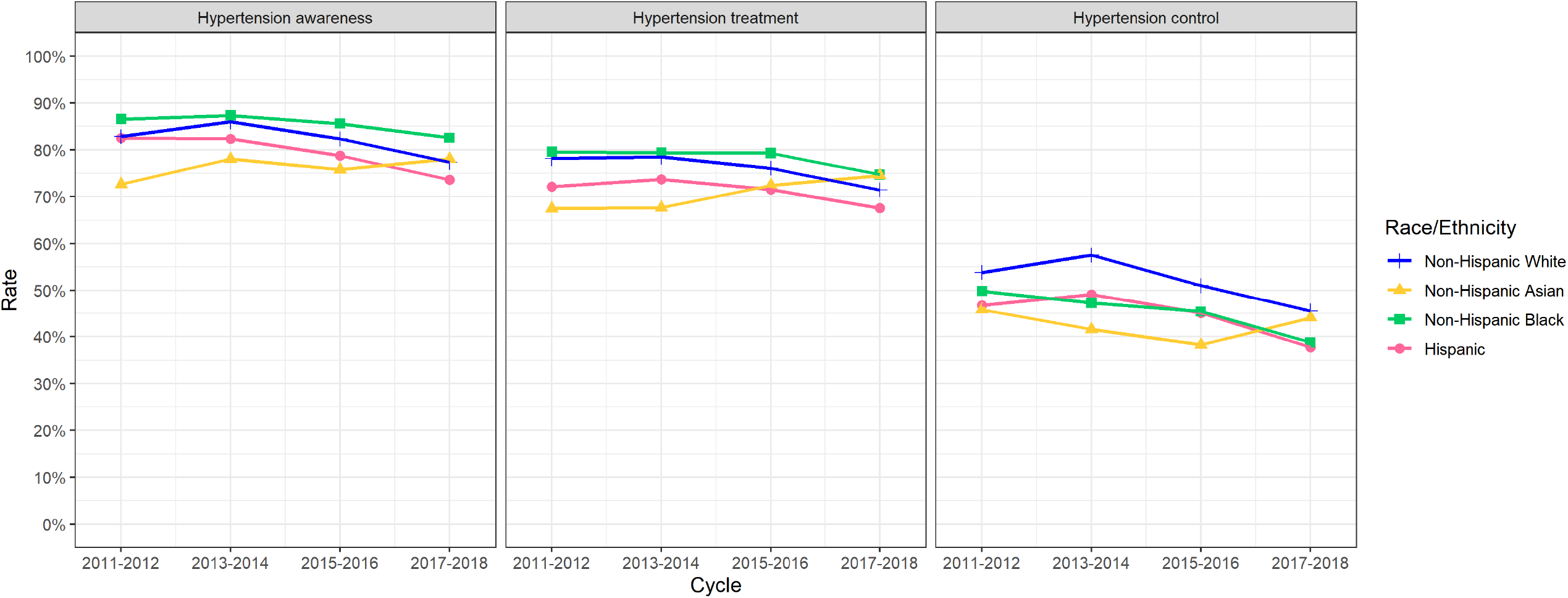
Racial and ethnic difference in awareness, treatment, and blood pressure control rate among all hypertensive adults, 2011-2018.

Compared with White individuals, Black individuals did not have a significantly different awareness rate (odds ratio:1.20 [0.96 to 1.45]), while Asian and Hispanic individuals had significantly lower rates (0.69 [0.52 to 0.85] and 0.74 [0.59 to 0.89], respectively) (Figure 4). Between 2011-2012 and 2017-2018, the racial/ethnic differences between White individuals and other groups did not change significantly (P>0.05 for all, Figure 1).

When stratified by age, sex, and income, the Asian-White and Hispanic-White differences in hypertension awareness persisted across all strata (Appendix Figure 2). The sensitivity analyses based on BP cutoffs in the 2017 ACC/AHA guideline provided consistent results (Appendix Figures 3-6).

### Trends in antihypertensive medication use by race and ethnicity

From 2011-2012 to 2017-2018, the age-adjusted treatment rate among hypertensive individuals declined from 77.3% (73.4 to 81.1) to 71.4% (69.0 to 73.8) in the overall population. This decline was consistent for Black (79.2% [75.5 to 82.9] vs. 74.7% [70.5 to 79.0]), Hispanic (71.7% [65.0 to 78.4] vs. 67.6% [62.5 to 72.7]) and White (78.3% [72.9 to 83.8] vs. 71.5% [67.5 to 75.0]) individuals. However, the treatment rate among Asian individuals improved significantly from 67.5% (61.9 to 73.1) to 74.5% (70.2 to 78.8) (Figure 1).

Among patients who received antihypertensive medication, the utilization of ACEIs, thiazide, and thiazide-like diuretics declined during the study period (Figure 2). However, there was an increase trend in utilization for ARBs, and no significant changes for β-blockers, CCBs, and other antihypertensive medications. Across all the drug classes, ACEIs were the most commonly utilized antihypertensive medication class accounting for more than 40% of all prescriptions throughout the study period.

**Figure 2.**
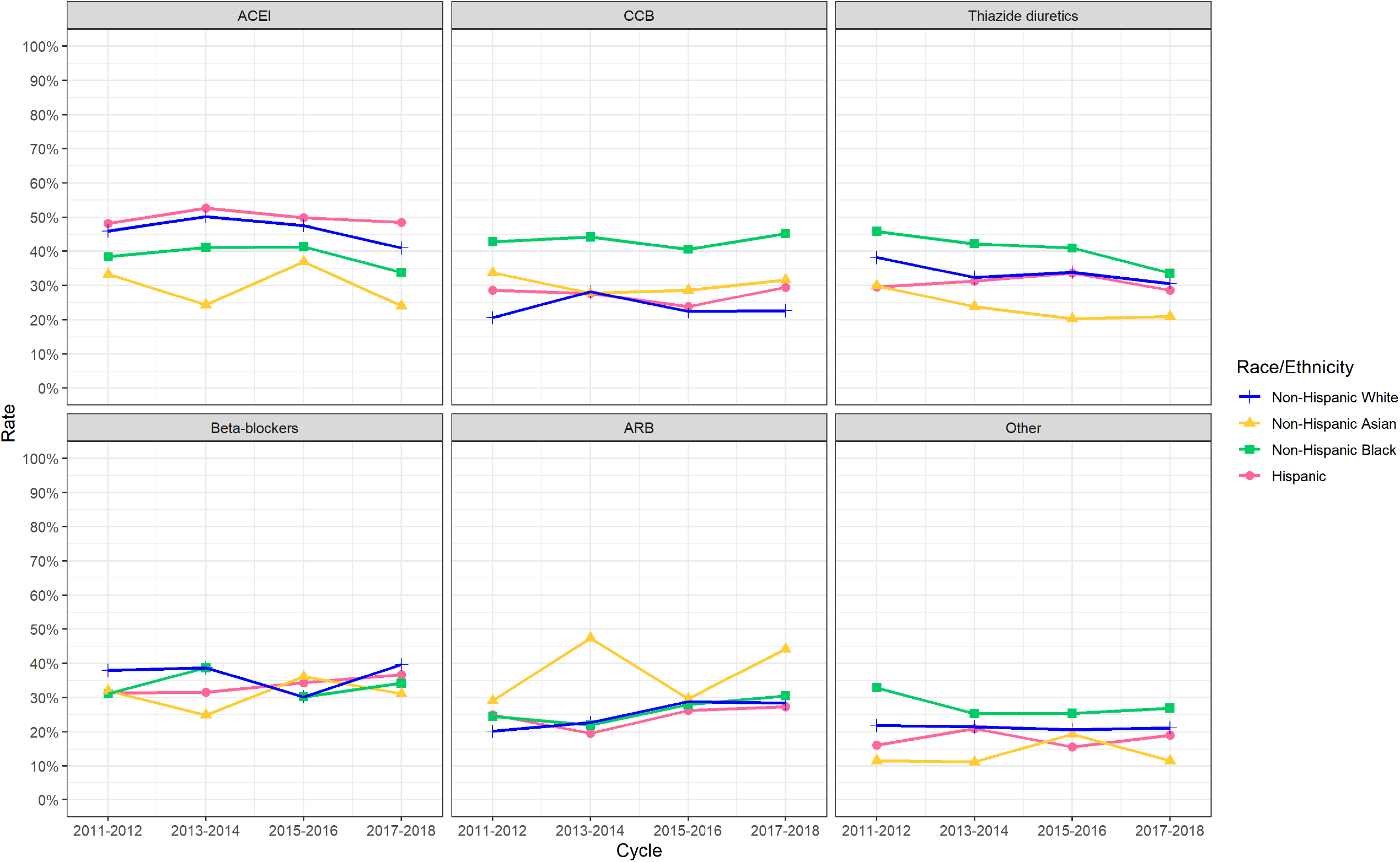
Racial and ethnic difference in type of antihypertensive medications among treated hypertensive adults, 2011–2018.

Compared with White individuals, Black individuals did not have a significantly different overall treatment rate (OR 1.04 [0.84 to 1.24]). However, when treated, they were significantly more likely to receive combination therapy (OR 1.45 [1.22 to 1.68]) and single-pill combination drugs (OR 1.36 [1.10 to 1.61]) (Figures 2-4). The utilization pattern for Black treated individuals was notable for significantly higher odds of receiving thiazide and thiazide-like diuretics (OR 1.29 [1.09 to 1.50]) and CCBs (OR 2.56 [2.24 to 2.89]) and lower odds of receiving β-blockers (OR 0.86 [0.74 to 0.98]) and ACEIs (OR 0.74 [0.62 to 0.87]) compared with White individuals. Unlike Black individuals, Asian and Hispanic individuals were significantly less likely to receive any antihypertensive therapy compared with White individuals (OR 0.72 [0.57 to 0.88] and 0.68 [0.55 to 0.81], respectively). Among those treated, Asian and Hispanic individuals received significantly fewer medications (OR for combination therapy, 0.60 [0.50 to 0.72] and 0.86 [0.75 to 0.96], respectively). Across all racial/ethnic subgroups, Asian individuals had the highest odds of receiving ARBs but the lowest odds of receiving ACEI and thiazide and thiazide-like diuretics. Between 2011-2012 and 2017-2018, the Asian-White differences in overall treatment rate significantly decreased over time due to the improvement in treatment rate among Asian individuals (P=0.05). The Hispanic-White differences in overall treatment rate also significantly decreased (P=0.03), but it was due to a larger decrease in treatment rate among Hispanic individuals. The treatment differences between Black and White individuals did not significantly change in this period.

**Figure 3.**
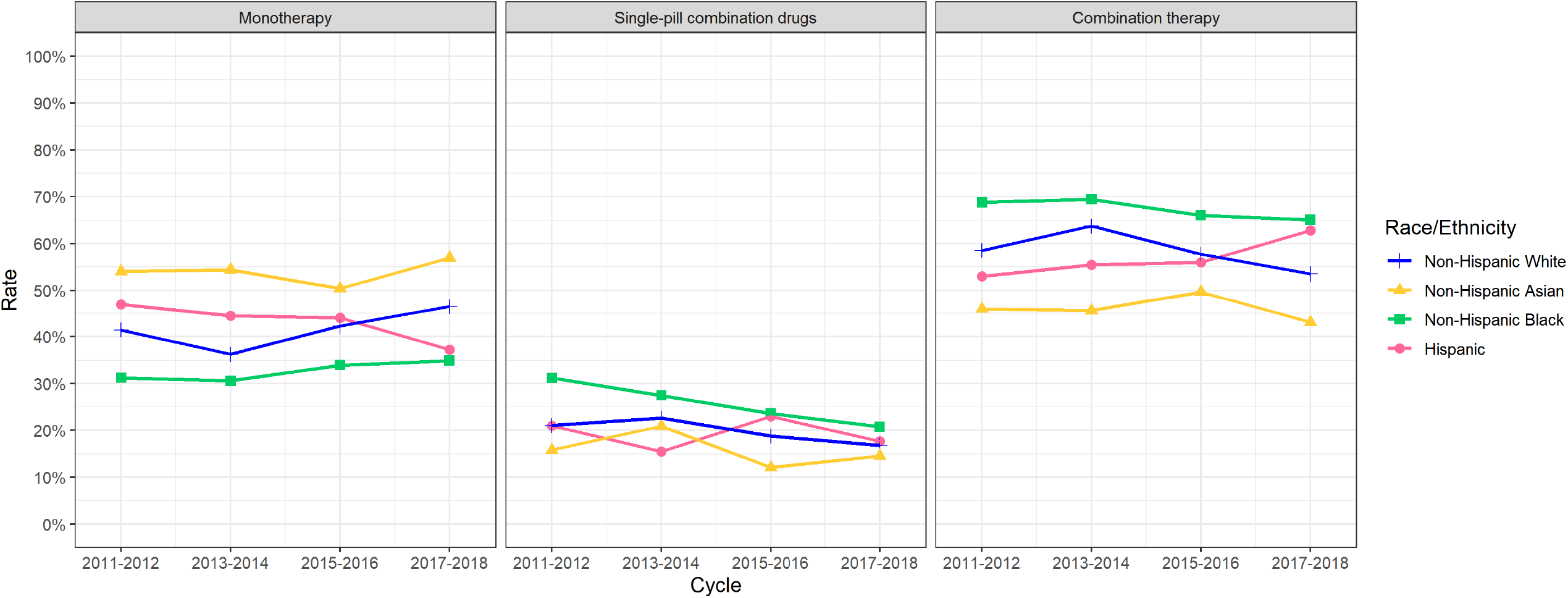
Racial and ethnic difference in number of antihypertensive medications among treated hypertensive adults, 2011–2018.

**Figure 4.**
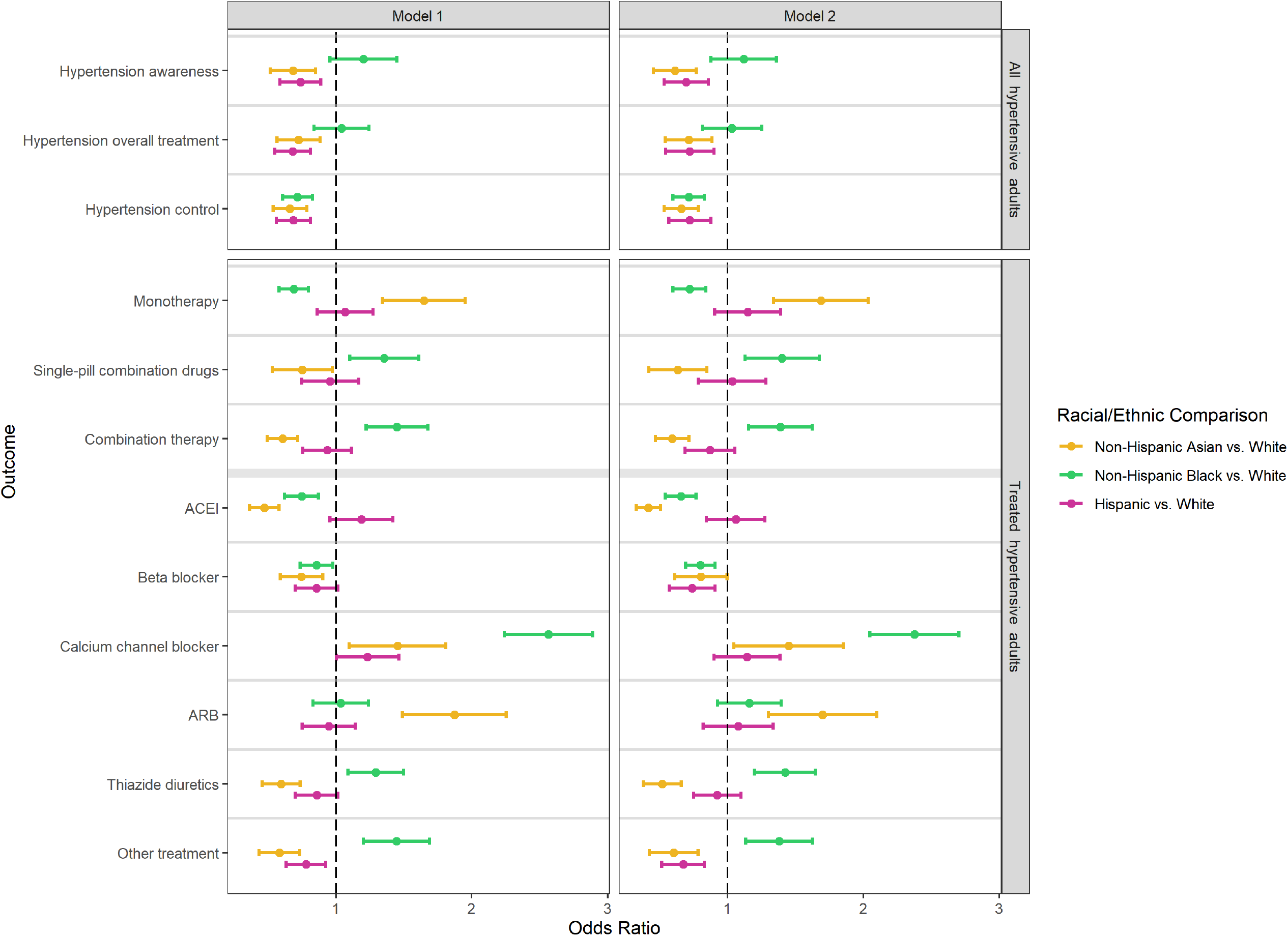
Odds ratios of racial/ethnic differences in hypertension awareness, treatment and control rates. Footnote: In model 1, we adjusted for participants’ age (categorized as 18-39, 40-59, 60-79, 80+ years of age) and sex. In model 2, we additionally adjusted for participants’ family income, education, insurance status, smoking status, diabetes, history of kidney diseases, and cardiovascular disease (MI or stroke).

When stratified by age, sex, and income, the racial/ethnic differences in hypertension treatment persisted across all strata. Asian and Hispanic individuals were less likely to receive any antihypertensive therapy and combination therapy compared with White individuals, while Black individuals had a similar overall treatment rate but were more likely to receive combination therapy (Appendix Figure 2). The sensitivity analyses based on BP cutoffs in the 2017 ACC/AHA guideline provided consistent results (Appendix Figures 3-6).

### Trends in BP control by race and ethnicity

From 2011-2012 to 2017-2018, the age-adjusted BP control rate among hypertensive individuals declined from 51.9% (47.1 to 56.7) to 43.1% (39.7 to 46.5) in the overall population. This decline was consistent in Black, Hispanic, and White individuals and was most obvious since 2013 to 2014. Specifically, the control rate declined from 49.8% (44.6 to 55.1) in 2013 to 2014 to 38.8% (33.8 to 43.8) in 2017-2018 among Black individuals, from 49.1% (43.6 to 54.7) to 37.8% (32.0 to 43.6) among Hispanic individuals, and from 57.5% (50.2 to 64.9) to 45.5% (40.6 to 50.4) among White individuals. If the control rates had not declined since 2013-2014, it was estimated that 986,111 more Black individuals, 1,084,653 more Hispanic individuals, and 6,240,265 more White individuals in the US would have been controlled in 2017-2018. Unlike other groups, the control rate among Asian individuals improved since 2015-2016 (38.3% [29.6 to 47.0] in 2015-2016 vs. 44.1% [38.9 to 49.3] in 2017-2018) (Figure 1).

Despite being similarly aware of hypertension and receiving more intensive therapy, Black individuals were less likely to achieve BP control (OR 0.72 [0.61 to 0.83]) compared with White individuals (Figure 4). The difference in BP control rates between White and Black individuals increased from 4.0 percentage points in 2011-2012 (53.8% vs. 49.8%) to 6.7 percentage points in 2017-2018 (45.5% vs. 38.8%), although the change was not statistically significant. Asian and Hispanic individuals were also less likely to attain the BP control goals compared with White individuals (0.66 [0.54 to 0.79] and 0.69 [0.56 to 0.81]). The difference in BP control rates between White and Asian individuals decreased from 8.1 percentage points in 2011-2012 (53.8% vs. 45.7%) to 1.4 percentage points in 2017-2018 (45.5% vs 44.1%). But the difference in BP control rates between White and Hispanic individuals did not change significantly, with a difference of 7.2 (53.8% vs. 46.6%) percentage points in 2011-2012 and 7.7 percentage points (45.5% vs. 37.8%) in 2017-2018.

When stratified by age, sex, and income, Asian, Black, and Hispanic individuals were less likely to achieve BP control compared with White individuals across all strata, and these racial/ethnic differences were more prominent in younger (<60 years of age), female, and lower to income individuals (Appendix Figure 1). The sensitivity analyses based on BP cutoffs in the 2017 ACC/AHA guideline provided lower control rates and persistent racial/ethnic disparities (Appendix Figures 3-6).

### Racial/ethnic differences in BP control and association with awareness and treatment

Appendix Figure 7 showed that balance was achieved after propensity score weighting. The weighted Asian-White and Hispanic-White differences in BP control were -5.2% (−12.5 to 2.1) and -8.5% (−15.0 to -2.0), respectively. They attenuated to -4.0% (−9.7 to 1.67) and 3.5% (−8.5 to 1.5) after additionally adjusting for hypertension awareness and treatment. On the other hand, the weighed Black-White difference in BP control rate was -6.6% (−11.2 to -1.9), and it was significantly magnified after additionally adjusting for hypertension awareness and treatment (−9.3 [-13.1 to -5.5]).

## DISCUSSION

In this nationally representative serial cross to sectional study, hypertension awareness, treatment, and control rates in the US were found to have plateaued or worsened from 2011-2018. Asian, Black, and Hispanic individuals had poorer hypertension control than did White individuals. Compared with White individuals, Black individuals had similar rates of awareness and treatment, and among those treated, they received a greater number of medications. Asian and Hispanic individuals had lower rates of awareness and treatment than White individuals, and among those treated, they received fewer medications. Racial/ethnic differences in awareness and treatment may partially explain the difference in hypertension control for Asian and Hispanic individuals, but not for Black individuals. These findings highlight the need for interventions to improve awareness and treatment among Asian and Hispanic individuals, and more investigation into the downstream factors that may contribute to the poor hypertension control among Black individuals.

Our study expands what is known about hypertension awareness and treatment in several ways. We showed that the previously described stagnant or decreasing hypertension awareness, treatment, and control in the US occurred in each of the racial/ethnic groups analyzed (except for Asians), with persistent racial/ethnic disparities. Such a lack of progress in hypertension control overall as well as reducing racial/ethnic disparities in hypertension management occurred during a period in which the total and per capita health care costs have increased substantially.^21 22^ Studies had reported similar race/ethnic disparities,^2 4 5^ but we analyzed yearly trends and racial/ethnic disparities in the intensity of treatment. Black individuals had higher rates of treatment and received more medications than White individuals, while Asian and Hispanic individuals had lower rates of treatment and among those treated, they received fewer medications. These racial/ethnic disparities did not significantly change from 2011 to 2018.

To the best of our knowledge, this is the first study to analyze recent national trends in hypertension awareness, treatment, and control among the Asian-American population and to compare these outcomes with those of other racial/ethnic subgroups. Previous studies on the Asian population covered a shorter time period and did not assess disparities with other racial/ethnic groups.^11 23 24^ During the study period, we found Asian individuals had significantly lower hypertension awareness, treatment, and control rates compared with White individuals, but all three metrics had improvements since 2015-2016. Notably, this is the only racial/ethnic subgroup that had improvement, while all other subgroups had worsening hypertension awareness, treatment, and control since 2015-2016. This finding signals the importance of investigating why the Asian population had improvement in hypertension management despite the slow progress nationally and identifying specific factors that can apply to other racial/ethnic groups.

This study has important public health implications. First, despite receiving more antihypertensive medications, Black people had a significant lower BP control rate compared with White people. There may be other responsible factors, including behavior, environmental, social and structural factors that disproportionally impair BP control among Black people. For example, Black people are more likely to have unhealthy dietary habits, sedentary lifestyle, poor sleep quality and stress compared with White people.^25^ They were also more likely to lack access to parks, healthy foods, and suffer from neighborhood noise.^26^ Additionally, as vast literature shows, systemic racism is a fundamental cause of health inequalities, impacting quality of care and the resources and opportunities available to support a healthy life.^27-29^ Therefore, efforts to increase hypertension awareness and treatment rates alone is insufficient to improve BP control among Black people. Public health strategies to improve hypertension control should be tailored to the needs of individuals and communities - this may include delivering care through trusted community resources,^30^ expanding the focus of BP interventions to include stress reduction and sleep quality, and working with policy officials to expand access to care and health resources. Second, the poorer hypertension control among Asian and Hispanic individuals is associated with their lower hypertension awareness and treatment compared with White individuals. Strategies to increase awareness and guideline to recommended antihypertensive medications are critical to improving hypertension control among Asian and Hispanic communities. Such strategies may include patient and provider education,^31 32^ home BP monitoring,^33^ behavioral counseling, and increase access to preventive care. These strategies can address misconceptions about hypertension,^34^ improve adherence to drug therapy,^35^ encourage lifestyle modifications, and improve access to care.

There are some differences between our findings and the results from other studies that have measured racial/ethnic differences in hypertension awareness, treatment, and control. First, the overall and race to specific hypertension awareness, treatment, and control rates in our study were lower than the estimates by Howard and colleagues, who analyzed data from the REasons for Geographic And Racial Differences in Stroke (REGARDS) study.^4^ This could be due to the different study population included in the analysis. The REGARDS study included participants > 45 years of age from a national cohort, while we included all participants ≥18 years of age from the NHANES national representative sample. Second, compared with Gu et al who also analyzed the NHANES datasets from 2003-2012,^5^ our study reports higher medication use and BP control rates among Black, Hispanic, and White individuals because of the inclusion of more recent data.

These findings are consistent with the positive trend seen in hypertension treatment and control from 2003-2012, following which treatment and control rates have either stagnated or decreased. Similar to their results, we report that Black individuals are still more likely to receive combination therapy but less likely to have BP control than White individuals, while Hispanic individuals continue to be under to treated and have lesser odds of having BP control than White individuals.

Our study has several limitations. First, the hypertension control targets changed over the study period. We used both the JNC 8 and 2017 ACC/AHA recommendations while understanding that neither of them is universally accepted. Second, the response rates in NHANES have declined over time. However, we used sampling weights developed by the National Center for Health Statistics to minimize nonresponse bias in our analyses. Third, the data on medication use was self to reported, therefore they may be subject to recall bias. Finally, this is a cross-sectional study with limitations attributable to the observational study design and residual confounding. The potential reasons for racial differences in hypertension control cannot be addressed by a single observational study, although we tried to assess the association between hypertension awareness, treatment, and control.

In conclusion, hypertension awareness, treatment, and control declined from 2011-2018 among adults in the US, and this decline was consistent for Black, Hispanic, and White individuals. BP control was worse for Asian, Black, and Hispanic individuals than for White individuals over the entire study period; this was explained partly by differences in awareness and treatment for Asian and Hispanic individuals, but not for Black individuals.

## Data Availability

Technical appendix, statistical code, and dataset available from the corresponding author.

## Copyright

The Corresponding Author has the right to grant on behalf of all authors and does grant on behalf of all authors, an exclusive license on a worldwide basis to the BMJ Publishing Group Ltd to permit this article (if accepted) to be published in BMJ editions and any other BMJPGL products and sublicenses such use and exploit all subsidiary rights.

## Details of contributors

All authors are responsible for study concept and design. YL and YLiu were responsible for the acquisition of data and its analysis. All Authors contributed to the interpretation of the data. YL drafted the manuscript. All authors performed critical revision of the manuscript for important intellectual content and approved the final manuscript. HMK is the guarantor.

## Ethical approval

Not required.

## Data sharing

Technical appendix, statistical code, and dataset available from the corresponding author.

## Transparency

The lead author affirms that this manuscript is an honest, accurate, and transparent account of the study being reported; that no important aspects of the study have been omitted; and that any discrepancies from the study as planned (and, if relevant, registered) have been explained.

## Patient involvement

No patients were asked for input in the creation of this article.

