## Supplemental material for "Trends in Awareness, Antihypertensive Medication Use and Blood Pressure Control by Race and Ethnicity Among Adults with Hypertension in the United States: A National Health and Nutrition Examination Analysis from 2011 to 2018"

**Online Supplement**

**Table of Contents**

| **Content** | **First Page** |
| --- | --- |
| **Appendix Table 1.** List of antihypertensive medications and classification | 2 |
| **Appendix Table 2.** Definition of sociodemographic, behavioral, and clinical variables in NHANES. | 3 |
| **Appendix Figure 1.** Study Population Flowchart | 5 |
| **Appendix Figure 2.** Racial/ethnic differences in hypertension awareness, treatment and control rates among all hypertensive adults, by age, sex, and income subgroups | 6 |
| **Appendix Figure 3.** Racial and ethnic difference in awareness, treatment, and blood pressure control rate among all hypertensive adults, using cutoffs in 2017 ACC/AHA guideline | 9 |
| **Appendix Figure 4.** Racial and ethnic difference in type of antihypertensive medications among treated hypertensive adults, using cutoffs in 2017 ACC/AHA guideline | 10 |
| **Appendix Figure 5.** Racial and ethnic difference in number of antihypertensive medications among treated hypertensive adults, using cutoffs in 2017 ACC/AHA guideline | 11 |
| **Appendix Figure 6.** Odds ratios of racial/ethnic differences in hypertension awareness, treatment and control rates, using cutoffs in 2017 ACC/AHA guideline | 12 |
| **Appendix Figure 7.** Balance assessment using Kolmogorov-Smirnov statistic | 13 |

**Appendix Table 1.** List of antihypertensive medications and classification

| **Drug class** | **Drug ingredient** |
| --- | --- |
| ACEI | Captopril, Enalapril, Fosinopril, Quinapril, Ramipril, Benazepril, Lisinopril, Moexipril, Trandolapril, Perindopril, Cilazapril |
| ARB | Losartan, Valsartan, Irbesartan, Eprosartan, Candesartan, Telmisartan, Olmesartan, Azilsartan |
| CCBs | Diltiazem, Verapamil, Nifedipine, Felodipine, Isradipine, Nicardipine, Nimodipine, Amlodipine, Nisoldipine |
| β-blockers | Atenolol, Labetalol, Nadolol, Propranolol, Acebutolol, Metoprolol, Pindolol, Timolol, Betaxolol, Penbutolol, Sotalol, Bisoprolol, Carvedilol, Nebivolol |
| Thiazide and Thiazide-like diuretics | Chlorothiazide, Chlorthalidone, Hydrochlorothiazide, Indapamide, Metolazone, Bendroflumethiazide, Methyclothiazide, Hydroflumethiazide, Trichlormethiazide, Hydrochlorothiazide, Polythiazide, Chlorthalidone |
| Others | Erythrityl Tetranitrate, Nylidrin Hydrochloride, Pentaerythritol Tetranitrate, Unspecified Antihypertensive Combinations, Clonidine, Furosemide, Diazoxide, Guanabenz, Guanethidine, Hydralazine, Methyldopa, Minoxidil, Prazosin, Acetazolamide, Amiloride, Bumetanide, Nitroglycerin, Reserpine, Spironolactone, Terazosin, Triamterene, Dichlorphenamide, Methazolamide, Ethacrynic Acid, Cyclandelate, Isoxsuprine, Papaverine, Ethaverine, Guanfacine, Guanadrel, Doxazosin, Alprostadil, Rauwolfia Serpentina, Torsemide, Spironolactone, Deserpidine, Tamsulosin, Alfuzosin, Eplerenone, Aliskiren, Silodosin, Riociguat, Sacubitril |

**Appendix Table 2.** Definition of sociodemographic, behavioral, and clinical variables in NHANES.

| **Risk factor** | **Ascertainment in NHANES** | **Definition of variables used in analysis** |
| --- | --- | --- |
| Marital status | In-person interview: Please describe your current marital status. | Marital status is classified as married/ living with partners vs. others (widowed, divorced, or separated; and never married). |
| Highest education level | In-person interview: What is the highest grade or level of school you have completed or the highest degree you have received? | Highest education level is classified as less than high school, high school, greater than high school. |
| Family income | In-person interview: Please describe your family income (reported as a range value in dollars) | Based on percent of family income relative to the federal poverty limit from the Census Bureau, family income is categorized as high/middle income [≥200%] and low-income [<200%] |
| Health insurance | In-person interview: Are you covered by health insurance or some other kind of health care plan? [Include health insurance obtained through employment or purchased directly as well as government programs like Medicare and Medicaid that provide medical care or help pay medical bills.] | Individuals are classified as insured if they had any private health insurance, Medicare, Medicaid, military plan, government or state-sponsored health plan. |
| Smoker status | In-person interview:   - Ever smoke cigarettes in entire life - Do you now smoke cigarettes? | Never smokers are defined as individuals who stated never smokes.  Former smokers are defined as individuals who stated that they had smoked but now did not smoke.  Current smokers are defined as individuals who stated that they smoked cigarettes currently. |
| Alcohol intake | In-person interview:   - Ever had a drink of any kind of alcohol - For the Past 12 months, how often do you have alcohol drink - For the past 12 months the average number of alcohol drinks per day | Never drinkers are defined as individuals who stated never had any kind of alcohol drink. Former drinkers are defined as individuals who stated they had had e drinks but did not drink for the past 12 months. Light drinkers are defined as individuals who self-reported 0-0.5 average daily drinks. Moderate drinkers are defined as individuals who self-reported (0.5-1.5 for women; 0.5-2.5 for men) average daily drinks. Heavy drinkers are defined as individuals who self-reported (1.5+ for women; 2.5+ for men) average daily drinks. |
| Physical activity | In-person interview:   - Behavioral Risk Factor Surveillance Survey (BRFSS) physical activity instrument. - Patients were asked if they participated in moderate or vigorous physical activity during the past 30 days. If they answered yes to either question, they were then asked the duration and frequency of their participation in physical activity for an average week. | Physical activity is classified into three levels: recommended (150 min or more of moderate physical activity or 75 min or more of vigorous physical activity per week), insufficient (between 10-149 min per week of moderate physical activity or between 10-74 min per week of vigorous physical activity), or inactive (no participation or fewer than 10 min of moderate or vigorous physical activity per week). |
| Obesity | All participants ages 2 and older had their standing height and weight measured during their physical examination. | Obesity is defined as body mass index (BMI)≥ 30kg/m^2^, where BMI is calculated as weight divided by the square of height. |
| Diabetes | In-person interview:   - Other than during pregnancy, have you ever been told by a doctor or health professional that you have diabetes or sugar diabetes? - Are you now taking diabetic pills to lower your blood sugar? These are sometimes called oral agents or oral hypoglycemic agents. - Are you now taking insulin?   Lab measurement: All participants ages 12 and older are given the option of a HbA1C% test during their physical examination | Diabetes is defined based on previous physician diagnosis, or HbA1c ≥ 6.5%, or currently on antidiabetic medication. |
| History of hyperlipidemia | In-person interview:   - Has a doctor or other health professional ever told you that you had high cholesterol? | History of hyperlipidemia is defined based on self-reported previous physician diagnosis of high cholesterol. |
| History of MI/stroke | In-person interview:   - Has a doctor or other health professional ever told you that you had heart attack? - Has a doctor or other health professional ever told you that you stroke? | History of MI/stroke is defined based on self-reported previous physician diagnosis of heart attack or stroke. |
| History of cancer | In-person interview:   - Has a doctor or other health professional ever told you that you had cancer? | History of cancer is defined based on self-reported previous physician diagnosis of cancer. |
| History of kidney disease | In-person interview:   - Has a doctor or other health professional ever told you that you had weak or failing kidneys? | History of kidney disease is defined based on self-reported previous physician diagnosis of kidney disease. |

**Appendix Figure 1.** Study Population Flowchart

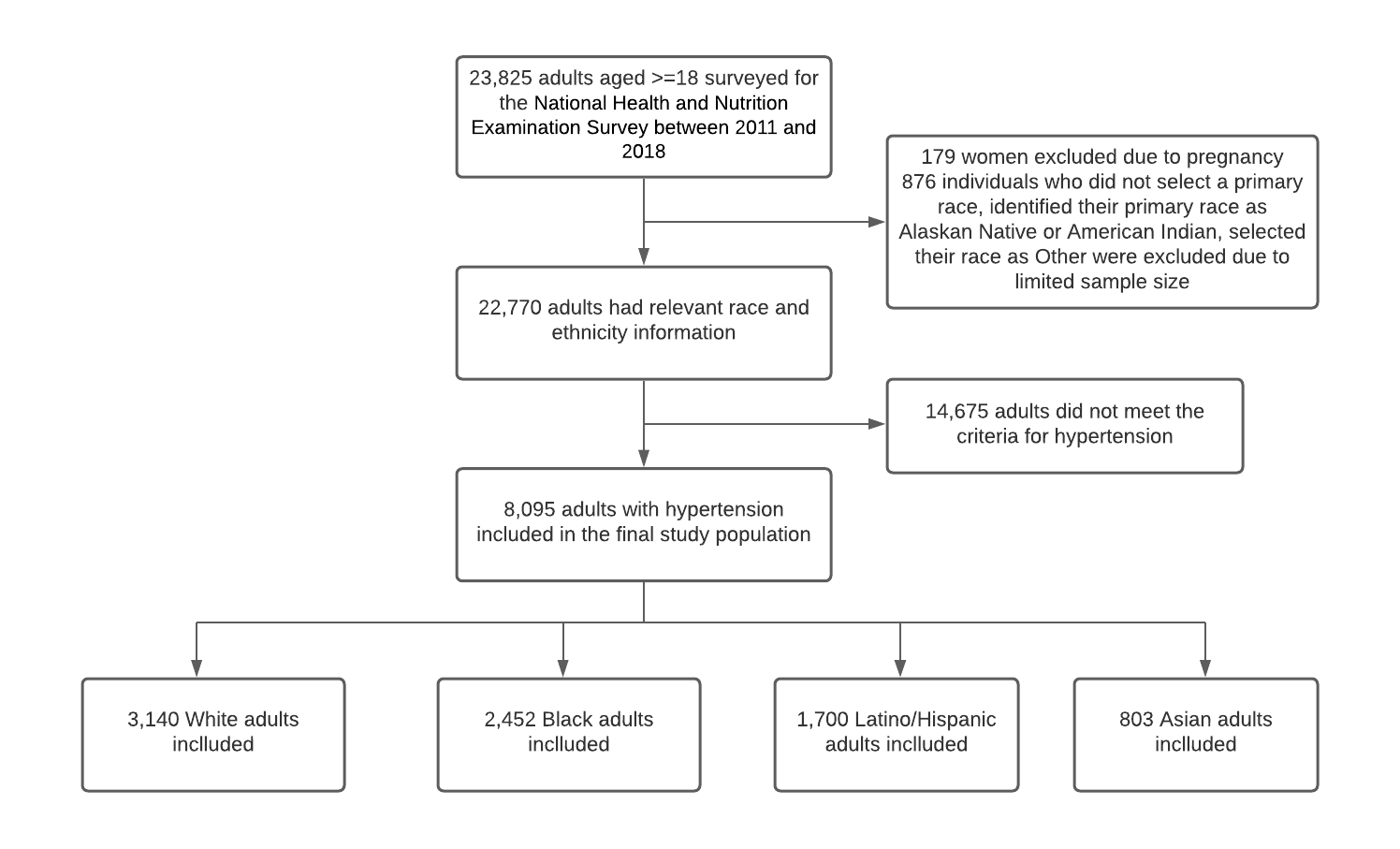

**Appendix Figure 2.** Racial/ethnic differences in hypertension awareness, treatment and control rates among all hypertensive adults, by age, sex, and income subgroups

**By Age (<60 vs. >=60)**

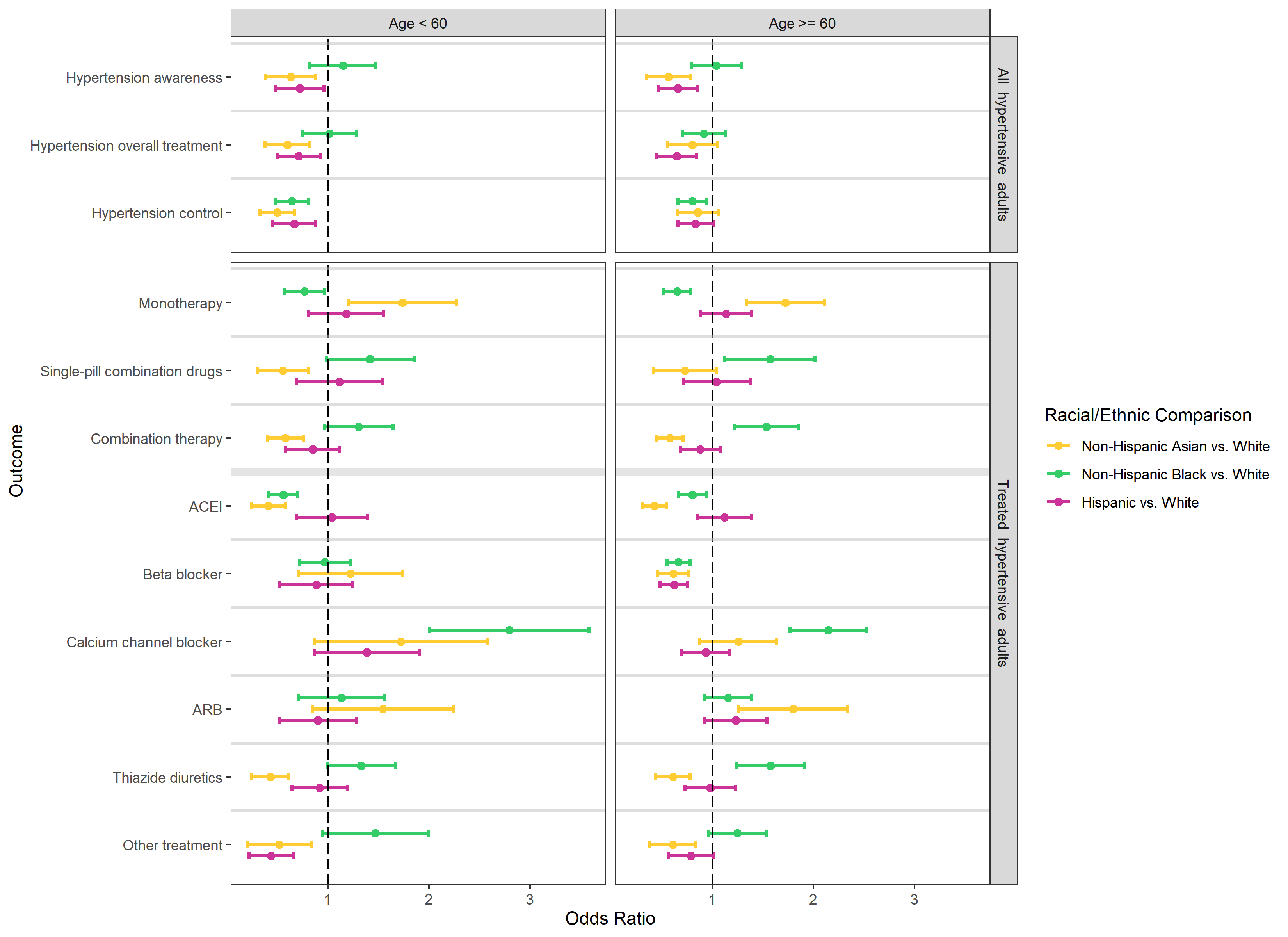

**By Sex (Male vs. Female)**

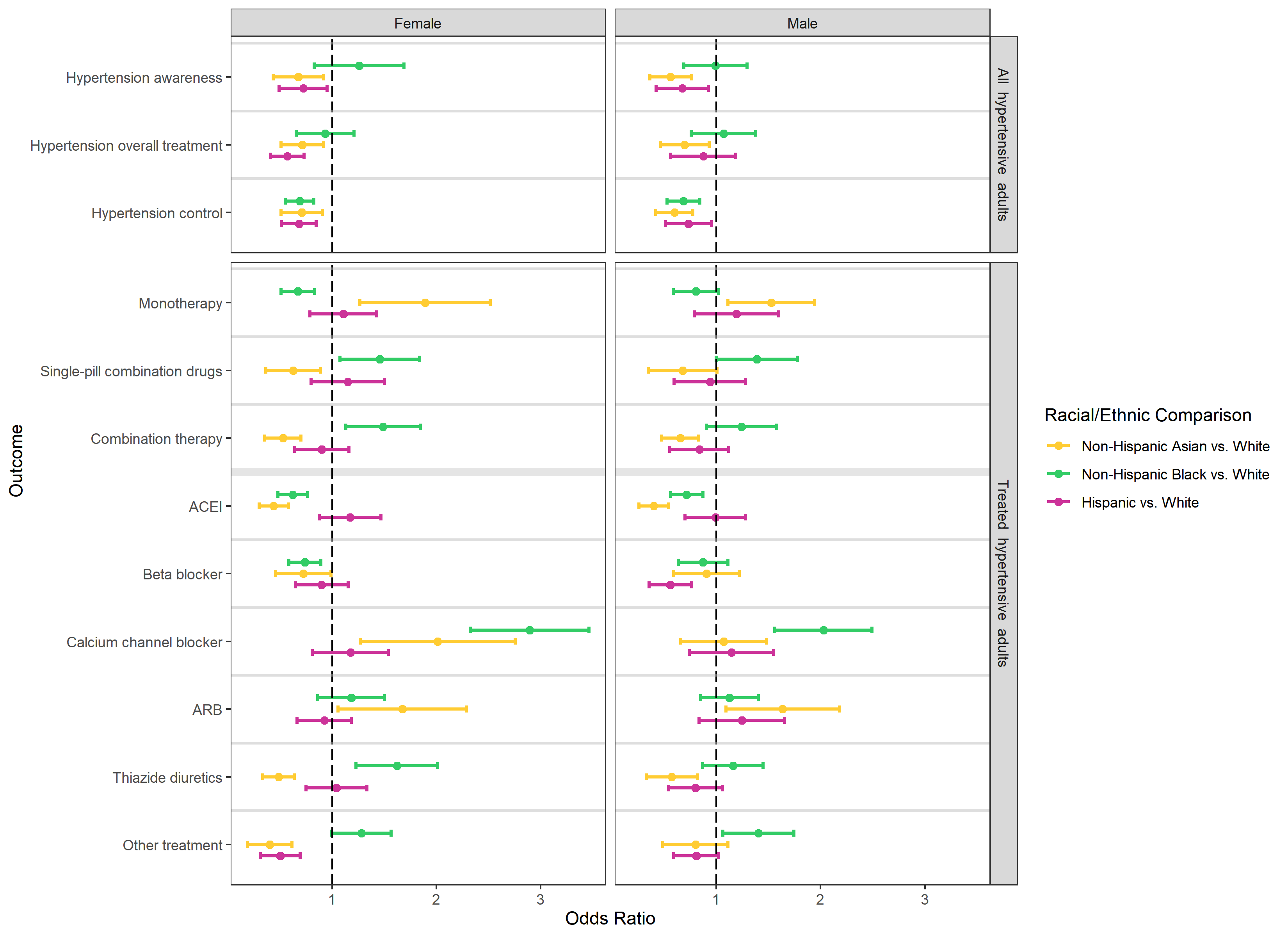

**By Income (high/medium vs. low)**

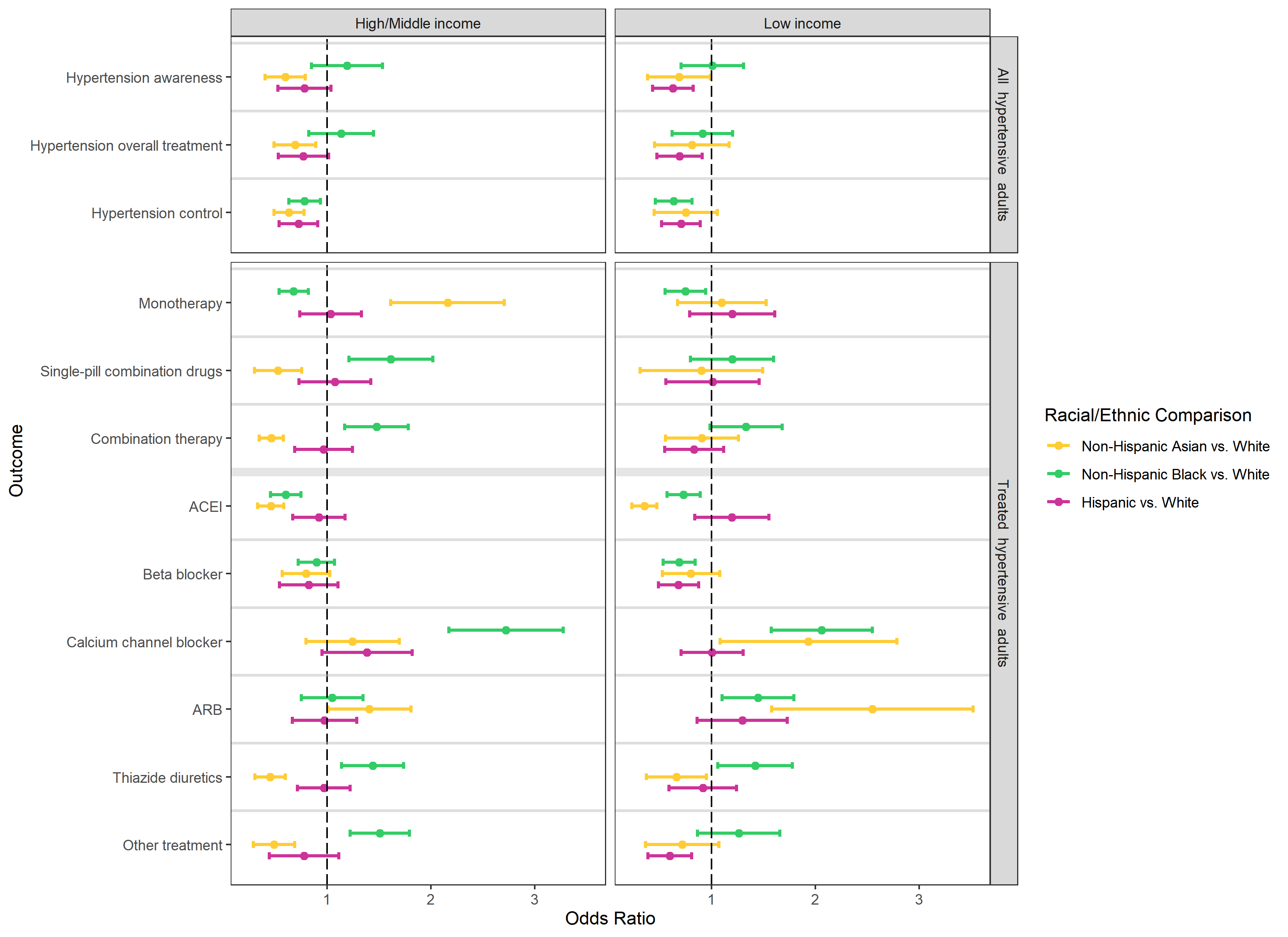

**Appendix Figure 3.** Racial and ethnic difference in awareness, treatment, and blood pressure control rate among all hypertensive adults, using cutoffs in 2017 ACC/AHA guideline

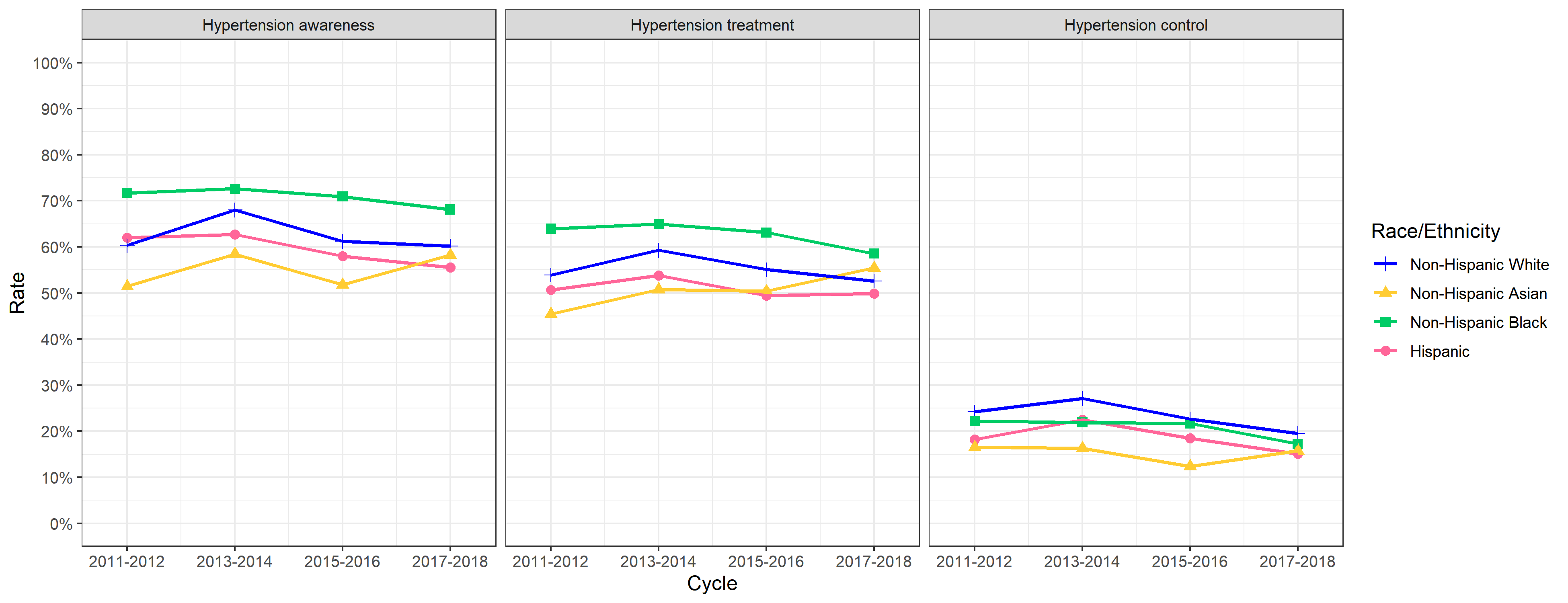

**Appendix Figure 4.** Racial and ethnic difference in type of antihypertensive medications among treated hypertensive adults, using cutoffs in 2017 ACC/AHA guideline

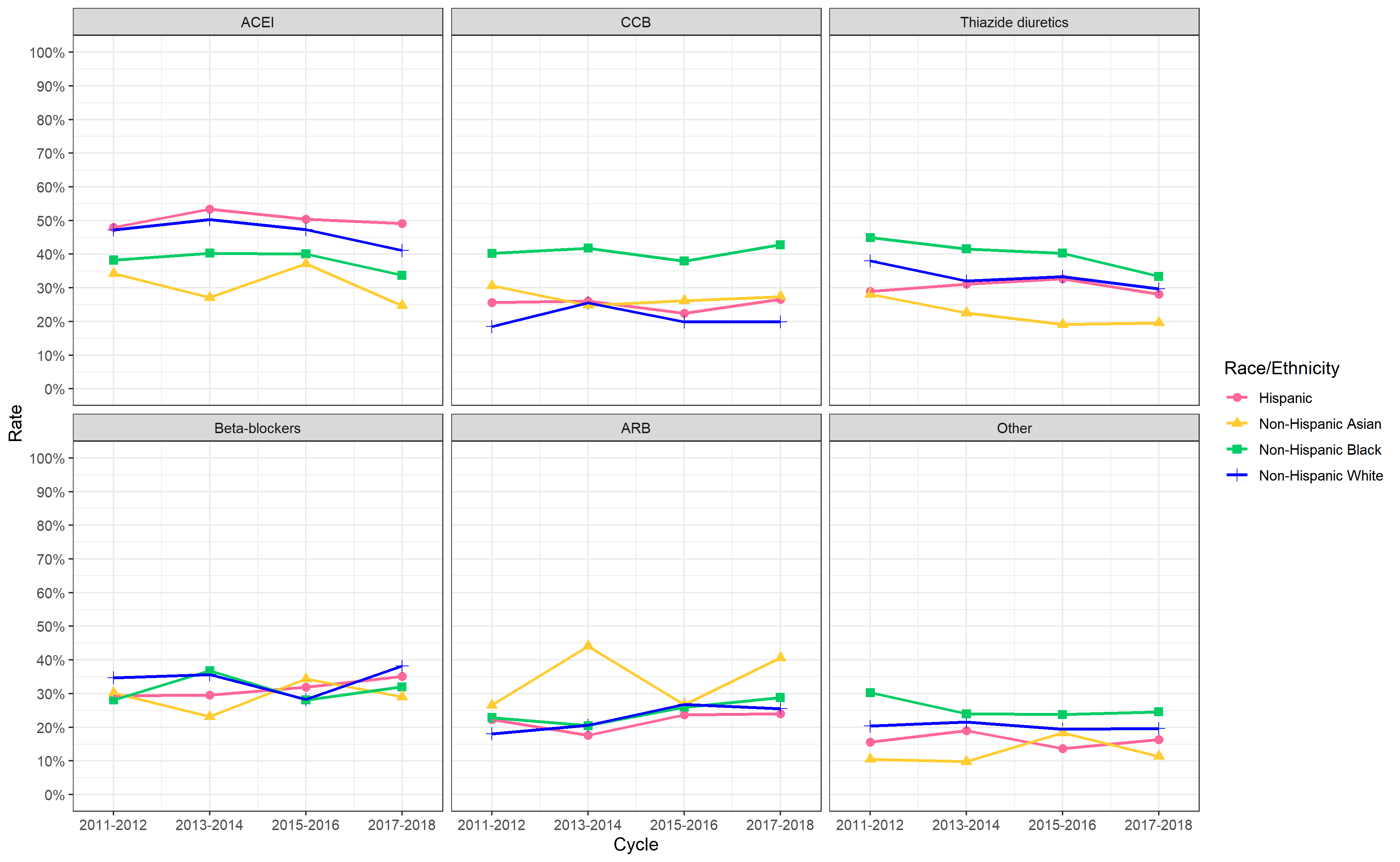

**Appendix Figure 5.** Racial and ethnic difference in number of antihypertensive medications among treated hypertensive adults, using cutoffs in 2017 ACC/AHA guideline

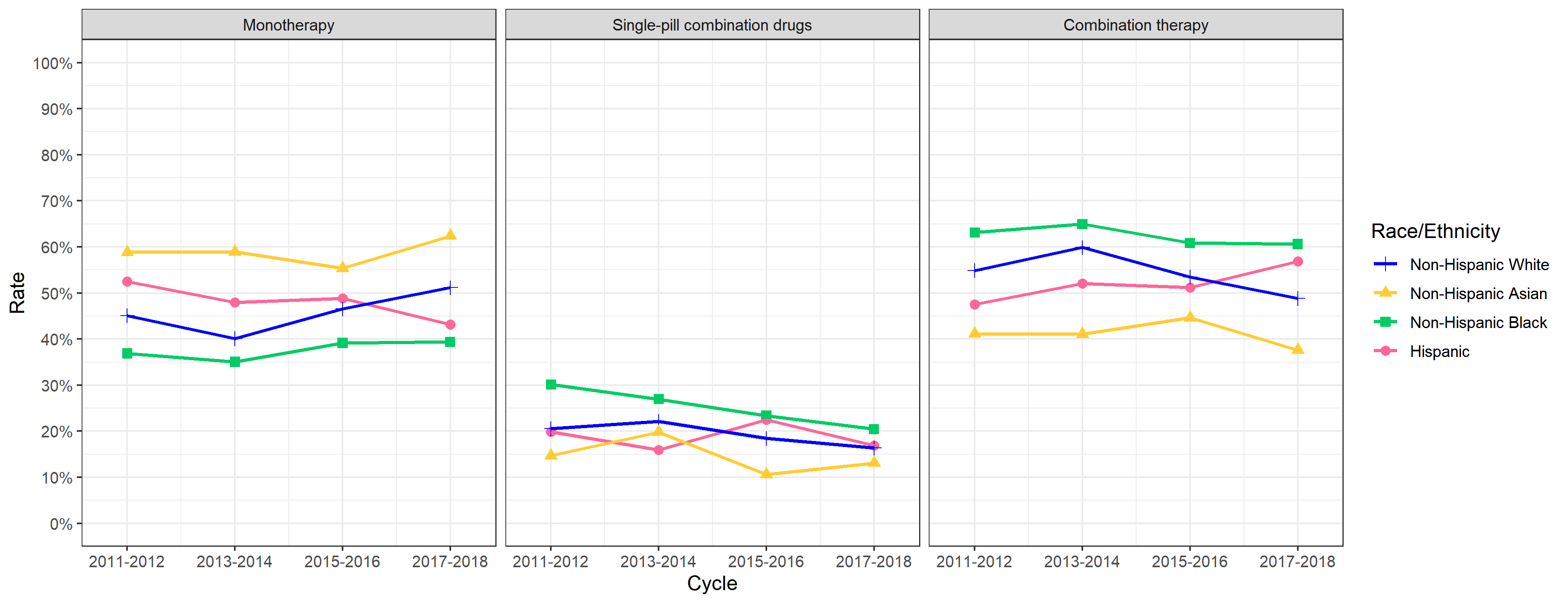

**Appendix Figure 6.** Odds ratios of racial/ethnic differences in hypertension awareness, treatment and control rates, using cutoffs in 2017 ACC/AHA guideline

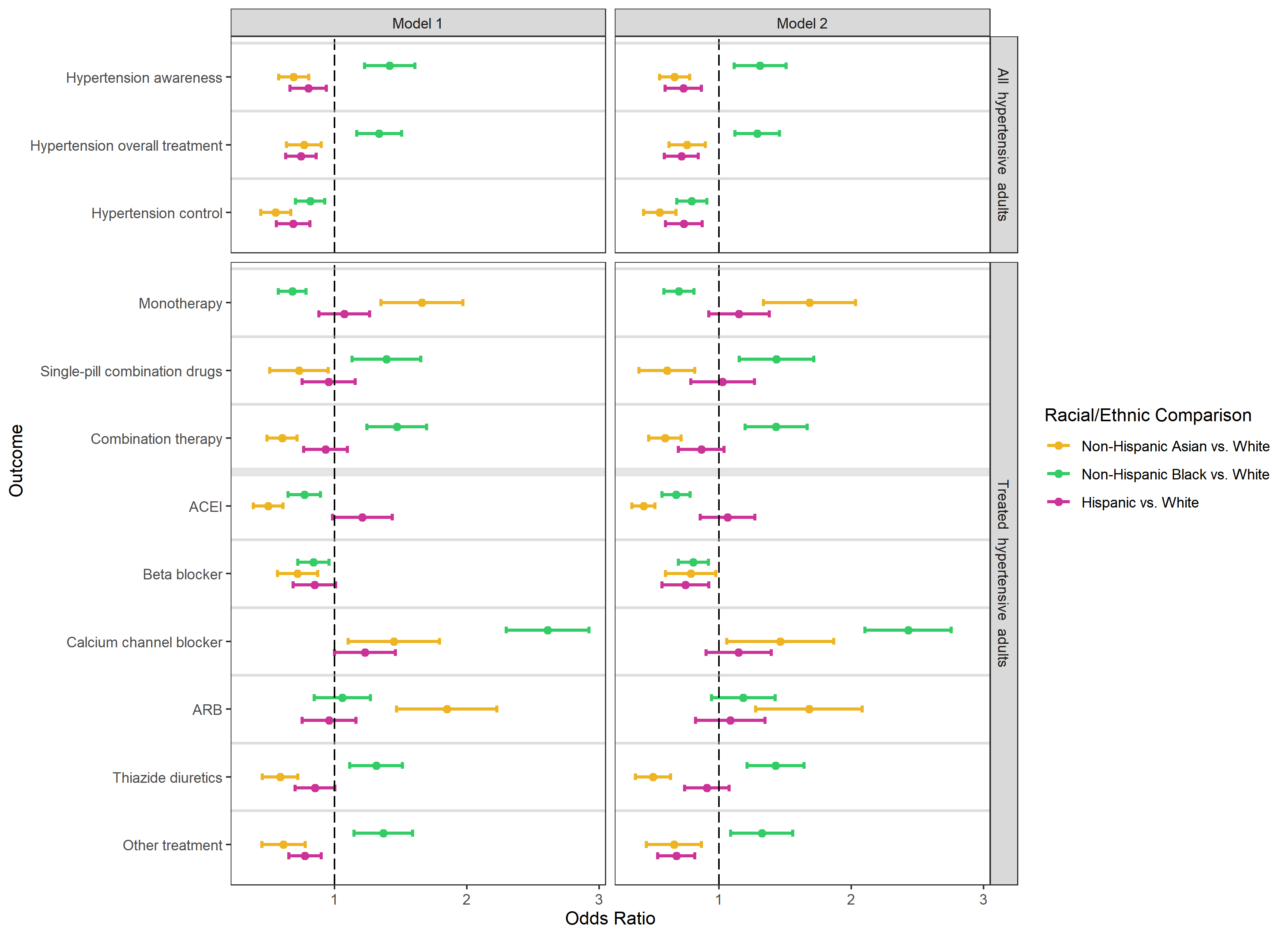

* In model 1, we adjusted for participants’ age (categorized as 18–39, 40–59, 60-79, 80+ years of age) and sex. In model 2, we additionally adjusted for participants’ family income, education, insurance status, smoking status, diabetes, history of kidney diseases, and cardiovascular disease (MI or stroke).

**Appendix Figure 7.** Balance assessment using Kolmogorov-Smirnov statistic

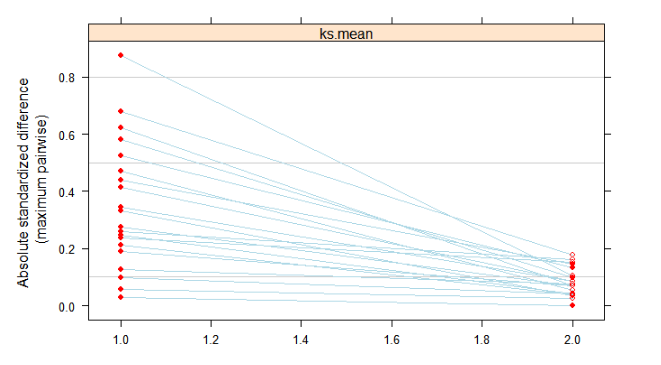

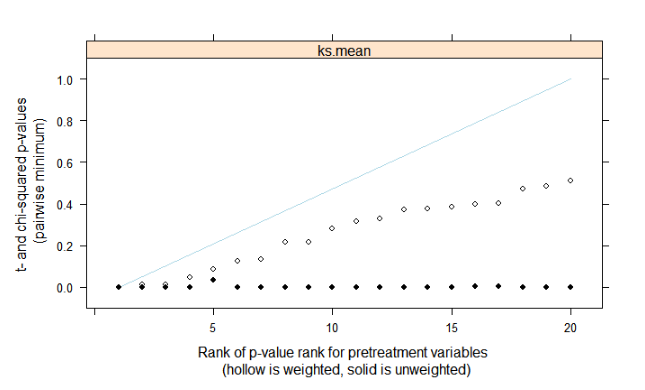
